## Supplementary File for "Using a real-world network to model the tradeoff between stay-at-home restriction, vaccination, social distancing and working hours on COVID-19 dynamics"

```
if X ≤ 80
    if X > 15
        if Z(t1 + t2) > Z(t3)
                Y classified as household contact
        else
                Y classified as workplace contact
        endif
    else
        if Z(t1 + t2) > Z(t3)
                Y classified as social environment contact
        else
                Y classified as workplace contact
        endif
    endif
endif
```

**Figure S1: Classification algorithm pseudocode.**

$Y$ represent encounters which occurred between 6m and 20m distance in Halsemere data point set, and $X$ represent total number of logged data points of each $Y$ during three consecutive day. The duration between 07:00 AM up to 08:30 AM and 18:00 PM up to 23:00 PM on Thursday and Friday represent by $t_1$. The duration between 07:00 AM up to 23:00 PM on Saturday represent by $t_2$. The duration between 08:30 AM up to 18:00 PM on Thursday and Friday represent by $t_3$. Let $Z(t_1 + t_2)$ give the total number $Y$ which occurred in $t_1$ and $t_2$ duration, and $Z(t_3)$ give the total number $Y$ which occurred in $t_3$

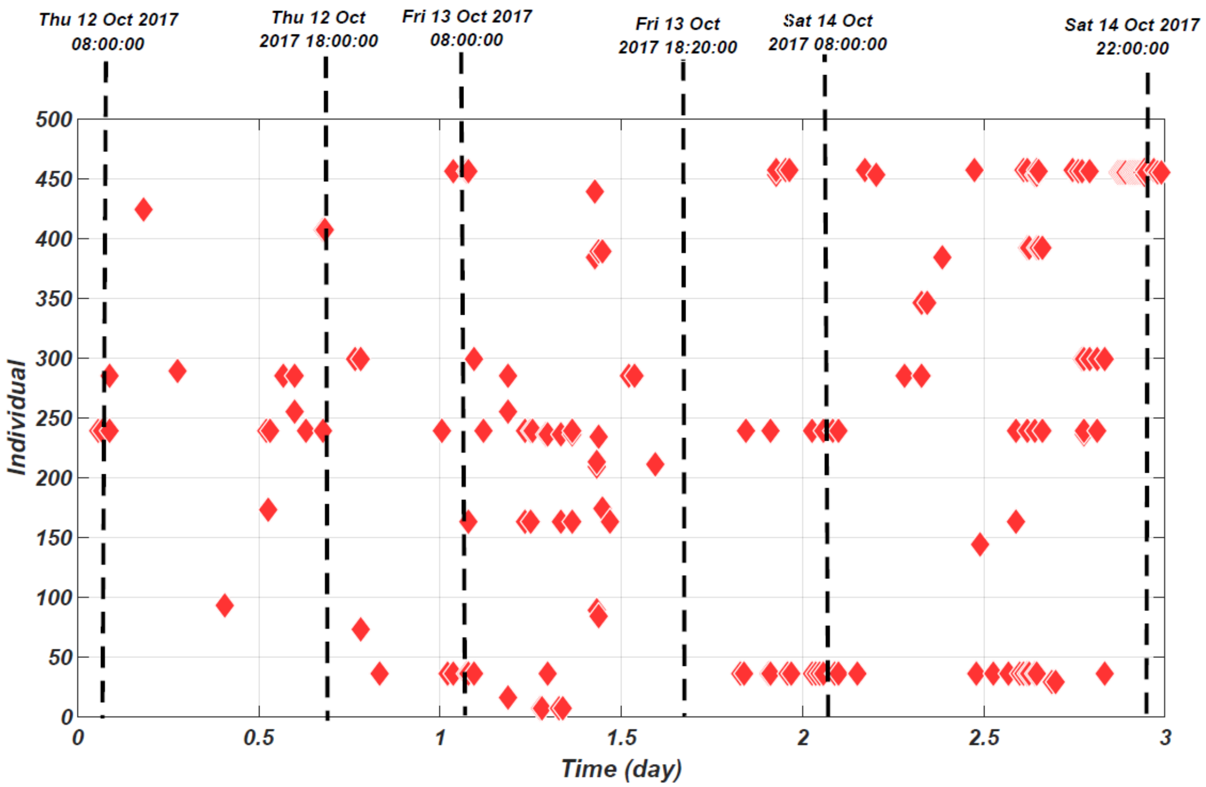


**Figure S2: Visualization method**.

Demonstrates the behavior and contact member of every single individual Haslemere data set during these three consecutive days. In here the behavior of 21st individual during three consecutive days. The red diamond shapes represent every single individual. The black dash line illustrates specific hours during these three days

**Table S1 list of procedures that are obtained by using visualization method**.

Demonstrates the behavior and contact member of every single individual of Haslemere data set during these three consecutive days

| Categories | Procedures |
|---|---|
| Household | 1) Individuals with at least 10 logged data points between 22:00 and 07:55 AM on any of the dates [1]<br>2) Individuals who contact for three consecutive night after 19:00 and have to distance less than 2<br>3) Individuals who have more than 80 logged data point during three consecutive day |
| Workplace | 1) Encounters which occurred on Thursday and Friday between 8:30 AM and 18:00 |
| Social Environment | 1) Individuals with a lease than ten logged data points between 22:00 and 07:55 AM on any of the dates<br>2) The encounters which are occurred after 19:00 clock for one-night lease than five logged data point<br>3) Encounters which occurred on Saturday from 8:00 AM and 18:00 less 15 logged data points |

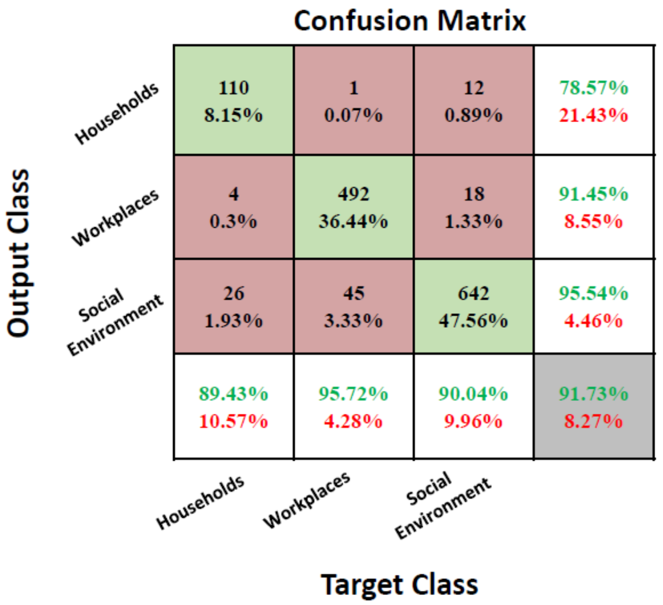


**Figure S3. Confusion matrix of classification algorithm for clustering Hazelmere data set into households, workplaces, and social environment with respect to visualization method classified data.**

In this figure, the first two diagonal cells show the number and percentage of correct classifications by the algorithm. For example, 110 contacts are correctly classified as household. This corresponds to 8.15% of all 1895 contacts. Similarly, 492 contacts are correctly classified as workplace. This corresponds to 36.44% of all contacts.in same way, 642 contacts are correctly classified as social environment. This corresponds to 47.56% of all contacts. 1 of the household contacts is incorrectly classified as workplace contact and this corresponds to 0.07% of all 1895 contacts. Also, 12 of the household contacts is incorrectly classified as social environment contact and this corresponds to 0.89% of all 1895 contacts. Similarly, 4 of the workplace contacts are incorrectly classified as households and this corresponds to 0.3% of all data. In same way, 18 of the workplace contacts are incorrectly classified as social environment and this corresponds to 1.33% of all data. 26 of the social environment contacts are incorrectly classified as households and this corresponds to 1.93% of all data. In same way, 45 of the social environment contacts are incorrectly classified as workplace and this corresponds to 3.33% of all data. Out of 123 household predictions, 78.57% are correct and 21.43% are wrong. Out of 514 workplace predictions, 91.45% are correct and 8.55% are wrong. Out of 713 social environment predictions, 95.54% and 4.46% are wrong. Out of 52 household contacts, 89.43% are correctly classified as household and 10.57% are classified as workplace and social environment. Out of 590 workplace contacts, 95.72% are correctly classified as household and 4.28% are classified as household and social environment. Out of 1003 social environment contacts, 90.04% are correctly classified as household and 9.96% are classified as workplace and households. Overall, 91.73% of the predictions are correct and 8.27% are wrong.

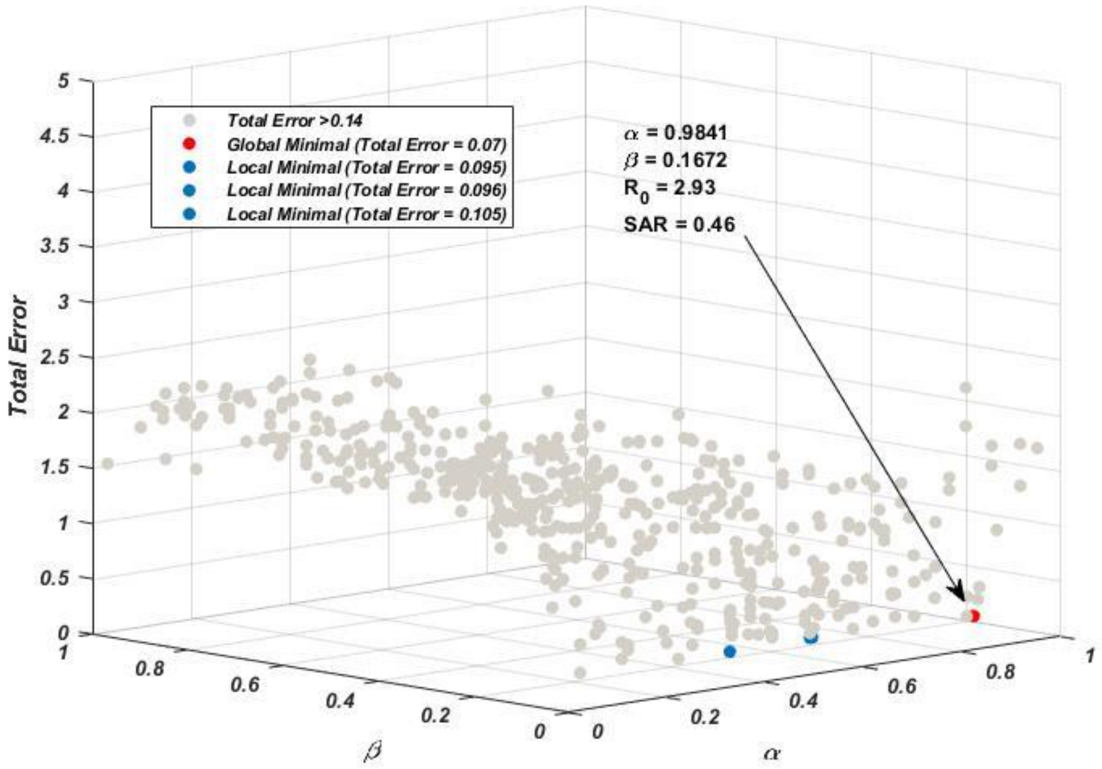


**Figure S4: Illustration of sampling result.**

For calculating scaling parameters of distance ($\boldsymbol{\alpha}$), and transmission rate ($\boldsymbol{\beta}$).

**Table S2 Agent based model parameters and their values**

| Parameter | Definition | Value | Source |
|---|---|---|---|
| $d_1$ | mean number of days in the latency stage | 2.7 day | Derived by finding the difference between incubation period and pre-symptomatic period. We take incubation period 5.1 day [7] |
| $d_2$ | mean number of days in pre-symptomatic stage | 2.4 day | [2] |
| $d_3$ | mean number of days in the asymptomatic stage | 5.4 day | Assumed to be the same duration as the total infectious period for symptomatic cases, including pre-symptomatic transmission [3] |
| $\boldsymbol{s}$ | the ratio of symptomatic cases | 0.83 | [4] |
| $d_4$ | average time of going to the hospital | 3 day | [5] |
| $\alpha$ | Infection Probability constant | 0.9841 | Derived (Fitted to COVID-19 $R_0$ and SAR of household by sampling (see SI Fig. 2)) |
| $\beta$ | Infection Probability constant | 0.1672 | Derived (Fitted to COVID-19 $R_0$ and SAR of household by sampling (see SI Fig. 2)) |
| $\mu_H$ | Transmission reduction fator for asymptomatic cases inside household | 0.696 | [6] |
| $\mu_O$ | Transmission reduction fator for asymptomatic cases outside household | 0.42 | [6] |
| $g$ | granularity | 192 | Derived by converting 1 day to 5 minutes intervals (16×12) |
